# Comparison of methods for identifying the optimal treatment duration in trials for antibiotics

**DOI:** 10.1101/2024.10.04.24314913

**Authors:** Suzanne M. Dufault, Brian H. Aldana, Patrick P.J. Phillips

**Affiliations:** Division of Biostatistics, Department of Epidemiology and Biostatistics, University of California, San Francisco, San Francisco, CA; Division of Pulmonary and Critical Care Medicine, Department of Medicine, University of California, San Francisco, San Francisco, CA

**Keywords:** tuberculosis, MCP-Mod, duration-ranging, antibiotics

## Abstract

**Background:** The optimal duration of antibiotic treatmentmust strike a delicate balance: it must be long enough to achieve desirable efficacy yet short enough to prevent the development of toxicities, adverse events, and mitigate other arduous aspects related to patient burden. Historically, the approach used to determine duration of antibiotic treatment has been inefficient, severely impacting the refinement of therapeutics for tuberculosis where treatment duration, and its complications, can be extensive. Meeting the WHO’s End TB target of developing shorter, more tolerable TB therapeutics will require rigorously re-evaluating best practices in duration-ranging trial designs and statistical methodologies. Many of the challenges in duration-ranging have parallels and proposed solutions in the field of dose-ranging where the literature is substantially more established and where the traditions of qualitative, pairwise comparison studies have been replaced with model-based approaches. Such methods are more efficient and allow for interpolation between the doses observed. Research on efficient study designs and methods for duration-ranging, while similarly attempting to capture a monotonic response relationship, has only just accelerated in earnest over the last two decades.

**Methods:** This work examines the utility of cutting-edge dose-finding methods (such as MCP-Mod) for duration-ranging of TB treatments. We compare the operating characteristics of the adapted model-based duration-ranging methodologies against standard qualitative methods for the purposes of estimating optimal duration and describing the duration-response relationship, using a simulation study motivated by a Multi-Arm Multi-Stage Response Over Continuous Intervention (MAMS-ROCI) clinical trial design. We explore three specific targets: 1) power to detect a duration-response relationship, 2) ability to accurately reproduce the duration-response curve, and 3) ability to estimate the optimal duration within an acceptable margin of error.

**Results:** We find that model-based methods outperform standard qualitative comparisons on every target examined, particularly when the sample size is constrained to that of a typical Phase II trial.

**Conclusions:** We conclude that the success of the next era in TB therapeutics duration evaluation trials, and antibiotics duration-ranging more broadly, will meaningfully rely on the ability to simultaneous pair innovative model-based statistical methods with re-imagined study designs such as MAMS-ROCI.

## 1 Introduction

Tuberculosis (TB) is a complex infectious disease requiring six months of multi-drug treatment with four drugs to effect cure; longer if the disease has resistance to key first-line drugs [1]. An estimated 10.6 million people developed TB in 2022, of whom only two thirds were diagnosed and started on treatment [1]. While the success rate hovers around 86% [2] when taken as intended, the many challenges associated with the arduous duration of treatment (e.g., toxicities, adverse events, patient burden) underscore the need for shorter, safer regimens [2, 3, 4] and many new drugs are in clinical development [5]. Duration of any new regimen is a critical yet delicate balance: it must be long enough to achieve desirable efficacy yet short enough to alleviate the aforementioned challenges.

Evaluating the optimal duration of a new regimen adds considerable complexity to clinical trials that must also identify the most favorable combinations of drugs at the proper safe and effective doses. Trials for new TB regimens have therefore typically evaluated only one or, at best, a narrow range of few durations within a single trial, comparing each shortened duration against a common control. Pairwise comparison of different treatment durations is not an efficient mechanism to determine the optimal duration, nor does it directly allow the exploration of intermediate durations that are not observed within the trial. The REMoxTB trial found two four-month regimens were not non-inferior to the six-month control [6], although mouse studies have since suggested that if one had been given for five months it probably would have achieved non-inferiority[7]. Bedaquiline was only studied for six months in clinical trials [8], but has since been show to be effective in longer regimens and WHO guidelines have included regimens with at least nine months of bedaquiline[9].

Many of the challenges in identifying an optimal duration have parallels and proposed solutions in trials seeking to identify an optimal dose, where the literature is substantially more established. Prior to the 1990s, dose-ranging studies approached dose as a qualitative nominal variable, evaluating the evidence for dose-response relationships using multiple comparison methods such as ANOVA and variations on Dunnett’s Method (a modified two-sample T-test approach) [10]. Such approaches do not permit interpolation or extrapolation of the dose-response relationship to dosages that were not studied and, as a result, tend to overestimate the minimum effective dose, increasing risks of toxicity, side effects, and other challenges to adherence. Approximately 10% of all drugs licensed by the FDA in the 1980s were recently re-dosed with most dosages dropped nearly 33% [10]. In the last few decades, model-based approaches for dose-finding have become more common. These methods treat observed dosages as observations on a continuous variable [11], are more efficient requiring fewer participants, and allow for extrapolation beyond and between the doses studied. Pre-specification of the corresponding dose-response curve is critical; some advocate the use of a fully flexible model (e.g., fractional polynomials, splines, nonparametric models) [12], others a slightly more restrictive monotonic flexible model (e.g., sigmoid Emax [13]), and still others advocate pre-specifying a candidate set of parametric models and a procedure for either model selection or model averaging [14]. This last approach, referred to as MCP-Mod, is a dose-finding method that combines the principles of multiple comparison procedures (“MCP”) with modeling (“Mod”). It has been shown to address the limitations of either approach performed in isolation and has been qualified by the USA Food and Drug Administration (2016) and the European Medicines Agency (2014) as an “adequate and appropriate method for dose selection” [15, 16]. The objective of our work was to adapt and evaluate best practice in the design and analysis of dose-ranging trials for use in duration-ranging trials.

## 2 Methods

The methods for this simulation study are organized according to the “Aims, Data-generating mechanisms, Methods, Estimands, Performance Measures” (ADEMP) framework proposed by Morris et al [17].

### 2.1 Aims

Two aims motivate this work. First, we aimed to adapt candidate model-based dose-ranging methodologies for the task of duration-ranging. Second, based on a range of expected duration-response profiles, we aimed to compare through simulation study the operating characteristics of model-based duration-ranging methodologies against standard qualitative methods to identify the preferred method for estimating the optimal duration and for describing the duration-response relationship.

### 2.2 Data-generating mechanism

In this simulation study we randomly assigned participants to the primary exposure of interest, *X*, duration of treatment, ranging from 8 to 16 weeks, evenly allocated across 2-week increments. This reflects the range of interest for drug-sensitive TB (DS-TB) therapeutics trials. The primary outcome, *Y*, was simulated as a binary endpoint defined as the presence of an unfavorable event (treatment failure or relapse) by the end of follow-up at 52 weeks post-randomization. The binary outcomes were simulated strictly as a function of treatment duration (*Y ~ f* (*X*)) according to a range of pre-specified duration-response curve shapes shown in Figure 1 (linear, log-linear, and logistic). Exact parameterizations are included in the Appendix (Table S1), and motivated by the expected efficacy of current DS-TB multidrug regimens [6].

**Figure 1:**
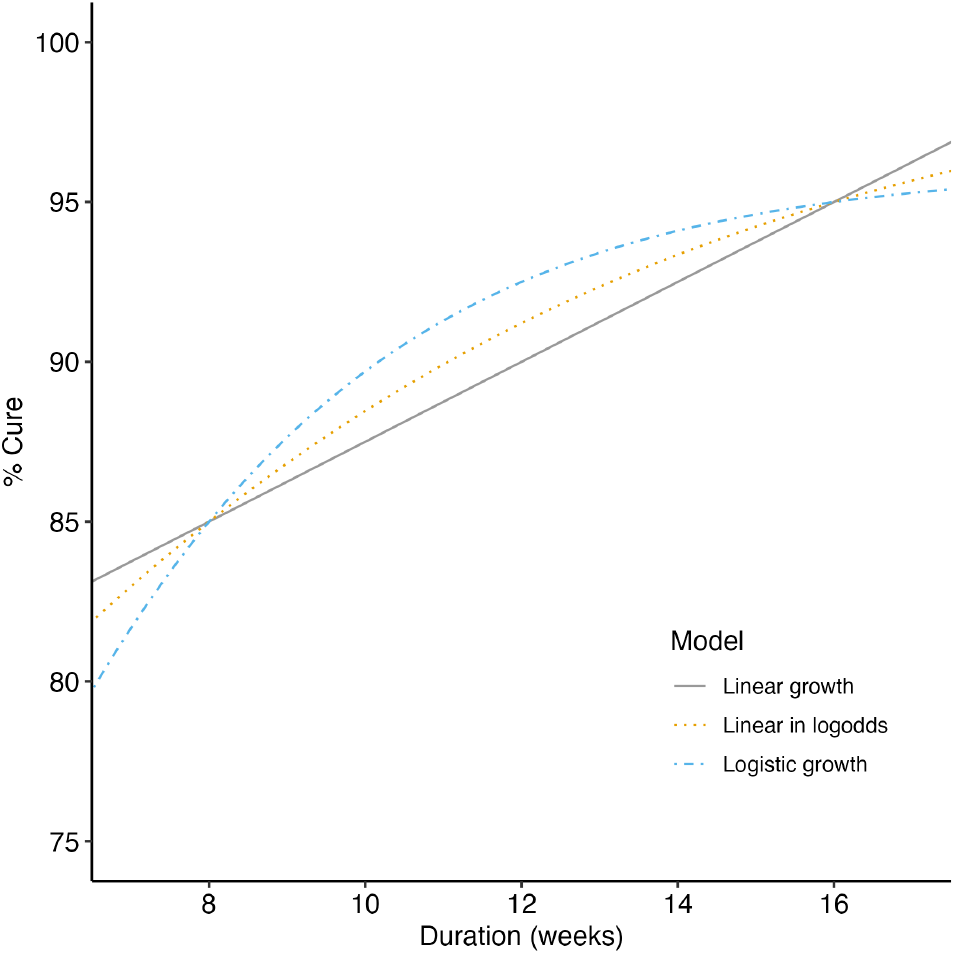
Data were generated according to parametric models. All models were parameterized such that 𝔼[*Y* |*X* = 8] = 0.85 and 𝔼[*Y* |*X* = 16] = 0.95. Exact parameter values are included in the Appendix (Table S1).

### 2.3 Estimands

The statistical targets (***ψ*** = {*ψ*_1_, *ψ*_2_, *ψ*_3_}) correspond to the duration-adapted motivating questions in Table 1. To target whether there is evidence of an effect (*ψ*_1_), we examined the statistical significance associated with testing the null hypothesis that there is no relationship between duration and response at an *a priori* specified *α* = 0.05. The second target, *ψ*_2_, is the estimated duration-response curve, and *ψ*_3_ is the estimated optimal duration, defined in this work as the minimum duration associated with a pre-specified “acceptable” relapse rate, specifically a relapse rate of 90%. We will refer to this as the minimum effective duration (MED).

**Table 1:** Motivating questions and statistical approaches – standard qualitative versus adapted model-based – used to estimate the corresponding targets. Note: optimal may be defined in many ways and the proposed approaches may not be appropriate for all definitions. For this work, we define optimal as the minimum duration associated with a pre-specified relapse rate.

| Question | Statistical Target | Standard Qualitative Approach | Adapted Model-based Approach |
| --- | --- | --- | --- |
| Is there evidence of an effect? | $\psi_1$ | Test of PoC: ANOVA | Test of non-zero slopes in candidate models |
| What is the duration-response relationship? | $\psi_2$ | – | Fitting of duration-response curve using candidate models |
| What is the optimal duration (weeks)? | $\psi_3$ | Pairwise one-sided tests of equivalence or non-inferiority, adjusted for multiple testing as appropriate | Direct estimation of optimal duration |

### 2.4 Methods of analysis

The statistical targets (***ψ***) were estimated using model-based and standard qualitative (pair-wise comparison) methods listed here (Table 2). The standard qualitative assessment framework does not directly empower estimation of the duration-response curve (*ψ*_2_), so only the model-based methods were compared on this target.

**Table 2:** Methods of analysis, their categorization as either novel model-based or standard qualitative, and the statistical targets (estimands) they can estimate.

| Method | Categorization | Targets Estimated |
| --- | --- | --- |
| MCP-Mod (select) | Model-based | $\psi_1, \psi_2, \psi_3$ |
| MCP-Mod (average) | Model-based | $\psi_1, \psi_2, \psi_3$ |
| Fixed 1-degree fractional polynomial | Model-based | $\psi_1, \psi_2, \psi_3$ |
| Fixed 2-degree fractional polynomial | Model-based | $\psi_1, \psi_2, \psi_3$ |
| Linear splines | Model-based | $\psi_1, \psi_2, \psi_3$ |
| Dunnett test | Qualitative | $\psi_1, \psi_3$ |

#### MCP-Mod

MCP-Mod can be utilized in either a model selection or model averaging framework; both were evaluated here. For both MCP-Mod procedures, the candidate model library included the following models, with parameterizations (Table S2) and visualizations (Fig S1) included in the Supplemental Material: Emax, sigmoid Emax, quadratic, and linear. For model selection (“MCP-Mod (select)”), the MCP-Mod procedure was performed accounting for the binary nature of the outcome variable. All candidate models in the candidate model library were fit. If at least one of the candidate models detected a significant relationship between duration and response, this was taken as evidence of effect (*ψ*_1_). The model with the “best” (i.e., minimum) gAIC was selected (MCP-step). This model was then used to estimate the duration-response curve (*ψ*_2_) and identify the MED (*ψ*_3_), defined as the minimum duration corresponding to an estimated mean response rate greater than or equal to 90% (Mod-step). For model averaging (“MCP-Mod (average)”), the MCP-Mod procedure was similarly performed for the MCP-step (*ψ*_1_). The Mod-step, however, was based on averaging the model fits from the “best” performing models across 100 bootstrap resamples of the data [11]. The model fits across the bootstrap resamples were then averaged in order to estimate the duration-response curve (*ψ*_2_), with the MED (*ψ*_3_) again defined as the minimum duration associated with a bootstrapped median response rate greater than or equal to 90%. The median of the bootstrapped response rates was chosen to be robust to outliers.

#### Fractional polynomials

Fixed 1- and 2-degree fractional polynomials were fit (“FP1” and “FP2”, respectively). To test whether a non-flat duration-response relationship exists, the gAIC of the fractional polynomial model was compared to the gAIC of an intercept only model. Evidence of effect (*ψ*_1_) was then determined when the fractional polynomial model had the lower gAIC. The fitted models were used to estimate the duration-response curve (*ψ*_2_) and to identify the minimum duration (*ψ*_3_) corresponding to an estimated mean response rate greater than or equal to a 90% response rate.

#### Linear splines

Piece-wise linear splines with 2 knots were fit under two conditions. First with equally spaced knots (LS2e) and second with 2 knots manually set at *k* = 10 and 12 weeks, as described in [4], concentrating the splines where the duration-response relationship is most likely to be non-linear. In both linear spline settings, evidence of effect was determined by a statistically significant, non-zero slope (*ψ*_1_). The fitted models were then used to estimate the duration-response curve (*ψ*_2_) and to identify the MED (*ψ*_3_) corresponding to an estimated mean response rate greater than or equal to 90%.

#### Dunnett test

A standard qualitative method for dose-ranging, we also performed Dunnett’s (1955) multiple comparisons procedure to compare the response rates among the discrete durations, using the longest duration as the control. Evidence of effect (*ψ*_1_) was established if there was at least one duration at which the observed relapse rate was estimated to be different from the longest duration’s relapse rate, at a statistically significant level (*P <* 0.05) where the p-value has been adjusted to account for multiple comparisons. Unlike the model-based methods, we are unable to pursue a direct estimate of the MED. Specifically, we cannot use the Dunnett method to directly identify the duration likely corresponding to a particular response rate. Instead, the Dunnett method does duration finding in terms of performance relative to the longest duration, which is expected to have the best performance in terms of proportion of cure. The MED (*ψ*_3_) was then determined one of two ways. First, we implemented a closed testing approach, performed as a step-wise procedure similar to that described in [10]. Beginning with the comparison between the longest and second longest durations, the estimated 90% simultaneous confidence bound on the difference in proportion of cure was compared to a clinically meaningful difference of 0.06. If the lower confidence bound was within this margin, the search for the optimal duration continued by comparing the results from the third longest duration versus those from the longest. This procedure continues until the lower confidence bound exceeds the margin, at which point the shorter duration has exceeded a clinically meaningful decline in performance relative to the longest duration available, and the penultimate duration evaluated is considered to be the MED. Second, we implemented an open testing approach, performed as an unordered procedure also described in [10], in which the MED (*ψ*_3_) was assumed to be the minimum duration whose 90% simultaneous confidence interval was estimated to fall below the clinically meaningful difference of 0.06, agnostic to any discontiguous duration-response intervals. In other words, the first approach requires that the duration compared against the “control” (i.e., the longest duration observed) was decreased in a step-wise, systematic way only when effectiveness is determined at each level. The second approach does not require finding evidence of step-wise effectiveness.

Exact parameterizations of each method can be found in the Supplemental Material (Section S2).

### 2.5 Performance measures

#### Power

Assuming each of the data-generating mechanisms were equally likely, the estimators’ performance in detecting evidence of effect (*ψ*_1_) was compared based on the proportion of simulated datasets across which there was sufficient evidence detected to reject the null hypothesis of a flat duration-response curve. The specifics of the test corresponding to each method are described in Section 2.4.

#### Scaled area between curves

The scaled area between the estimated and true curves [4] was used to reflect estimator performance in capturing the true duration-response curve (*ψ*_2_). This metric averaged the absolute model performance across the entire curve and was scaled such that the possible values range from 0 to 1, where 0 is a perfect fit. Qualitative approaches cannot estimate the duration-response curve (*ψ*_2_), and therefore performance on this target was only compared among the modeling methods.

#### POS_MED_

While there were three targets of interest, priority was given to the estimation of the optimal duration (*ψ*_3_), defined as the minimum duration associated with a relapse rate of 90%, or MED. The statistical performance of each estimation method in this regard was measured as the probability of successfully identifying the true optimal duration within a certain range (POS_MED_) [18]. This performance metric accounts for both estimator bias (i.e., accuracy in estimating the true optimal duration) and variance (i.e., the expected precision associated with the estimator) when evaluating estimator performance.

Assuming each of the data-generating mechanisms were equally likely, we examined POS_MED_ under three conditions, outlined in Eq. 1-3. First, we examined the probability that the estimated MED from a given method fell within the interval between the true MED and the maximum duration observed (Eq. 1). This is aligned with what Quartagno et al [19] refer to as the “acceptable power” which targets “finding an effective duration that is shorter than the recommended one, even if it is not necessarily the *minimum* effective duration.” Next, we examined the probability that the estimated minimum duration fell within two intervals around the true minimum where both intervals assume it is better to err on selecting a duration that is slightly too long than one that is too short. We examined the probability that the estimated MED was no shorter than the truth by one (Eq. 2) or two weeks (Eq. 3), while also being no more than two (Eq. 2 or four (Eq. 3) weeks longer than the truth. As an example, if the true optimal duration (*ψ*_3_) is 9 weeks, Eq. 2 would accept as “successful” any method with an estimated duration 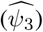 ranging between 8 and 11 weeks while Eq. 3 would expand the definition of success to any estimated duration 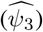 ranging between 7 and 13 weeks.

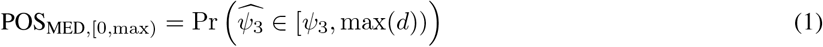

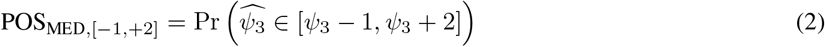

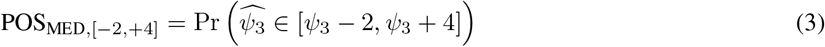

The performance of all three metrics for varying sample sizes was used to explore estimator robustness.

All statistical estimation methods, model-based and qualitative, were ranked on the basis of their performance; the most desirable regimens having high power, low scaled area between the estimated and true curves, and a high POS_MED_. All code necessary for data generation, analysis, and report compilation is publicly available in a GitHub repository maintained by the first author (https://github.com/sdufault15/tb-mcp-mod). All analyses were performed in R version R 4.4.1 (2024-06-14) “Race for Your Life”, using ‘DoseFinding’[20] and ‘MCPMod’[21] packages for performing the MCP-Mod analyses, ‘lspline’[22] for the linear splines, ‘gamlss’[23] for the fractional polynomials, ‘DescTools’[24] for the Dunnett Tests, ‘tidyverse’[25] and ‘patchwork’[26] for data wrangling and figure generation.

## 3 Results

### 3.1 Method Performance Overall

A high-level summary of each method’s performance across various metrics is shown in Table 3. These results are compiled for 15,000 simulated datasets at each sample size, assuming each data-generating mechanism is equally likely. Performance across each data-generating mechanism individually is explored in the subsequent subsections. The best performance for each metric at each sample size is highlighted in gray for ease of comparison.

**Table 3:**
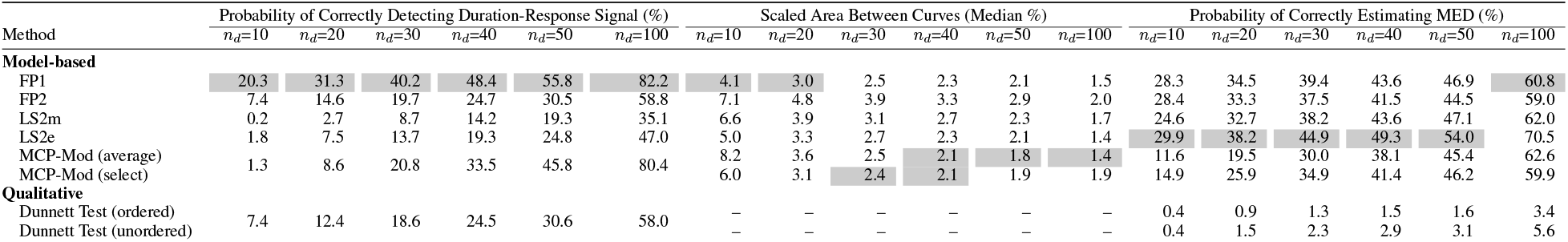
Probability of correctly detecting a duration-response signal, probability the estimated minimum effective duration (MED) falls within a reasonable range around the true MED (POS_MED,[−1,+2]_), and the scaled area between the true duration-response curve and the estimated duration response curve by sample size for each estimation method. The best performing method at each sample size for each metric is highlighted in gray.

The power to detect evidence of effect (*ψ*_1_) is captured in the first set of columns. As expected, across each method, the performance in detecting a non-flat duration-response relationship increases as the sample size randomized to each duration (*n*_*d*_) increases. Note, the two MCP-Mod approaches apply the same method to detect evidence of effect (MCP-step), and therefore, their results are identical and pooled in Table 3. The same is true for the two Dunnett Test approaches; their results have been similarly pooled for presentation. Fractional polynomials have the best performance in detecting a duration-response signal, with 20% probability when there 10 participants per duration (*n*_*total*_ = 50). The other methods, model-based and Dunnett, require approximately 30 participants per duration (*n*_*total*_ = 150) to achieve the same probability of correctly identifying evidence of effect when it is present.

The method-specific performance in terms of accurately estimating the underlying duration-response curve (*ψ*_2_) summarized via the scaled area between curves metric is captured in the second set of columns. Here, the MCP-Mod methods have the best performance with only 2.4% average absolute error in estimating the probability of cure across the range of durations observed when there are 30 participants randomized to each duration (*n*_*total*_ = 150). FP1 is similarly competitive across the range of sample sizes as well.

Method-specific performance in accurately estimating the MED (*ψ*_3_) within a reasonable range around the truth is captured in the third set of columns. Specifically, these columns capture the proportion of analyses where the estimated MED fell within an interval of no less than one week below the true MED and no less than two weeks greater than the true MED (POS_MED,[−1,+2]_). Given the relatively linearity of the underlying DGMs, it is perhaps unsurprising that the linear spline methods tended to have best performance in terms of estimating the MED within the acceptable interval, despite having poor overall fit of the duration-response curve. The Dunnett Test has the poorest performance, consistently failing to select any duration lower than 16 weeks (Supplemental Figure S2).

### 3.2 Performance: Evidence of Effect

As seen in Figure 1, the data have been generated from a particularly narrow range of expected effect sizes, allowing for a maximum difference in proportion of cure of 10% (85% efficacy to 95% efficacy). The FP1 and MCP-Mod methods achieved at least 80% power when 100 participants were randomized to each duration. Across sample sizes less than 100 per duration, the estimated power was lower than would be acceptable in a classic Phase III study, but may be sufficient for an exploratory Phase II study. For example, the maximum probability of detecting evidence of an effect only reaches 58.8%, even at a sample size of 50 per duration (Figure 2).

**Figure 2:**
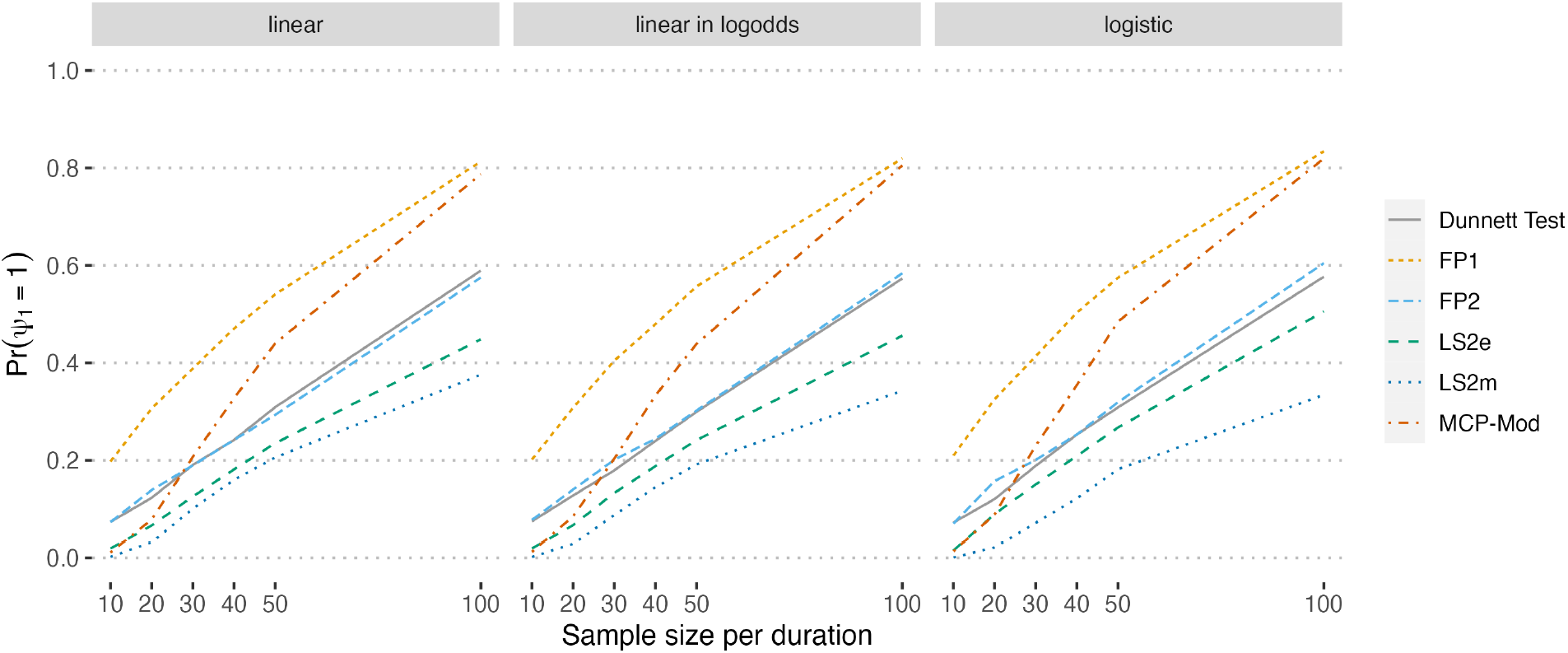
The proportion of simulated datasets across which there was sufficient evidence detected to reject the null hypothesis of a flat duration-response curve (power).

The methods perform quite similarly in terms of power across all data-generating mechanisms, with slightly improved performance observed in the logistic growth setting (Figure 2: logistic). The FP1 method dominates across the range of sample sizes considered, though the MCP-Mod approach grows increasingly competitive as the sample size increases. The power to detect an effect for the Dunnett Test is not notably different than the other model-based methods and has the second highest power when the sample size is extremely small (*n*_*d*_ = 10).

### 3.3 Performance: Duration-Response Curve Fit

Estimated duration-response curves from the model-based fits to the data generated by the logistic data-generating mechanism (solid black line) with a sample size of 30 participants per duration (150 total) are shown in Figure 3. For this sample size and data-generating mechanism, it is visually apparent that the linear spline models are parameterized in ways that force an over-complicated estimated shape. The fractional polynomial models tend to fare better, though FP2’s tendency to under- and overestimate the response rates corresponding to lower durations is concerning. Overestimation of the response rate, in particular, could lead to falsely selecting a MED that is considerably lower than the true optimal duration required to achieve a target response rate. The MCP-Mod methods both appear to generally mimic the proper shape, with a slightly lower, though non-negligible risk of rate overestimation at lower durations and occasional risk of incorrectly suggesting a non-monotonic relationship.

**Figure 3:**
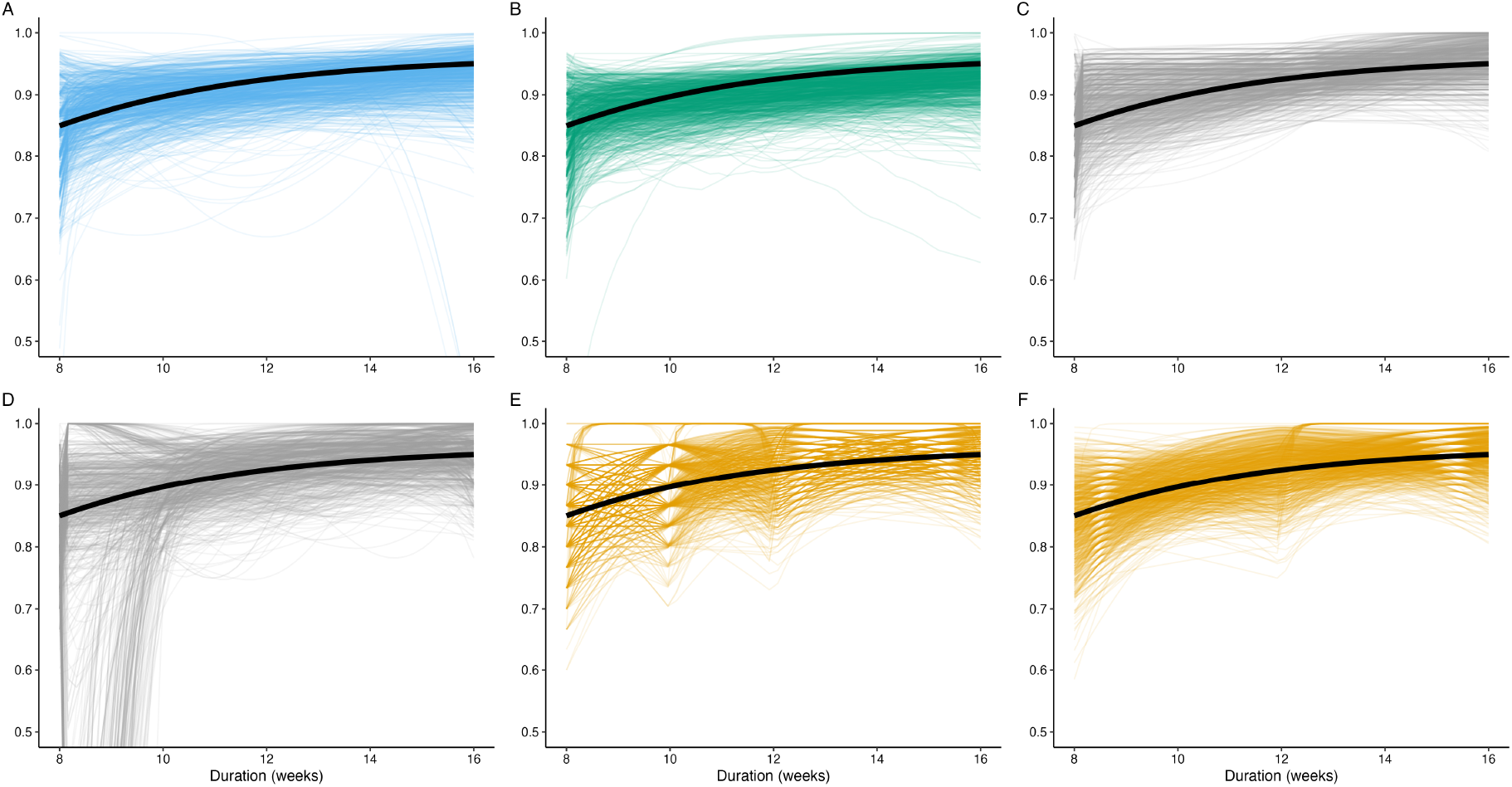
Estimated duration-response curves from 1,000 datasets simulated from the logistic data-generating mechanism plotted against the true duration-response curve (black) for a sample size of 30 participants per duration, evenly distributed across the five durations. A) MCP-Mod (select), B) MCP-Mod (average), C) FP1, D) FP2, E) LS2m, F) LS2e.

To quantify the model-based performance in estimating the duration-response curves, we examine the results of the scaled area between the curves analysis (Figure 4). Only the results for the logistic data-generating mechanism are shown here, though results across all data-generating mechanisms were similar and can be found in the Supplemental Material (Figure SS3). The scaled area between the curves metric captures the average absolute error in estimating the probability of cure across the whole duration range, scaled to fall between 0 and 1, and reported here as a percentage. Figure 4 shows the median average absolute error (%) for each method across the simulated datasets. The results from this analysis confirm what was observed in Figure 3. All methods are quite accurate at large sample sizes, with a maximum average absolute error of less than 2.5% when 100 participants are assigned to each duration (*n*_*total*_ = 500). As sample size decreases, FP2 tends to have the highest error in estimating the probability of cure across the whole range of durations considered. FP1, on the other hand, is consistently competitive with the MCP-Mod approaches in terms of lowest average absolute error.

**Figure 4:**
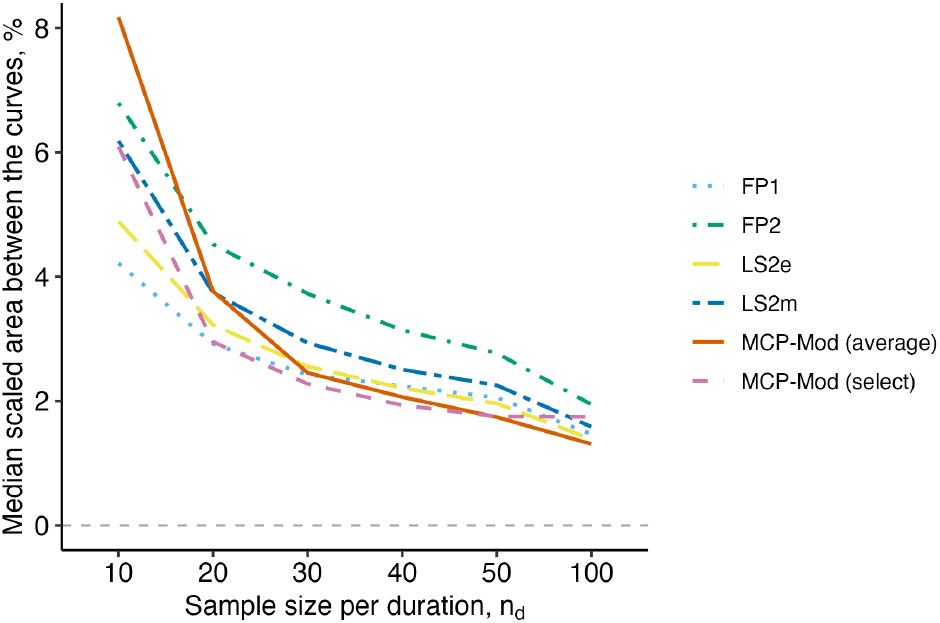
The median scaled area between the curves (%) from the estimated fits from 5,000 simulated datasets for each method, by sample size assigned to each duration (*n*_*d*_). The data were simulated based on the logistic data-generating mechanism. A scaled area between the curves (%) of zero would indicate a perfect fit.

For further insight, Figure 5 shows the estimated duration-response curves associated with the worst model fits (largest scaled area between the curves) for sample sizes of 10, 30, 50, and 100 per duration. FP2 (Figure 5D) and the linear spline with manually placed knots (Figure 5E) have erratic worst fits that appear to run the risk of overestimating the effectiveness associated with shorter durations. This could lead to decreasing the duration beyond the MED. FP1 (Figure 5C) and the linear spline with equally placed knots (Figure 5F) have worst fits that are desirably monotonic and conservative in their estimation of the duration-response relationship. Finally, the difference in the MCP-Mod (select) and MCP-Mod (average) methods (Figure 5A,B) is well illustrated here. When using MCP-Mod (select) with a small sample size, there is a risk of performing estimation based on an incorrect model. This risk is somewhat mitigated when MCP-Mod averages across the best model fits. This performance may further improve for MCP-Mod (average) with the application of an increased number of bootstrap samples.

**Figure 5:**
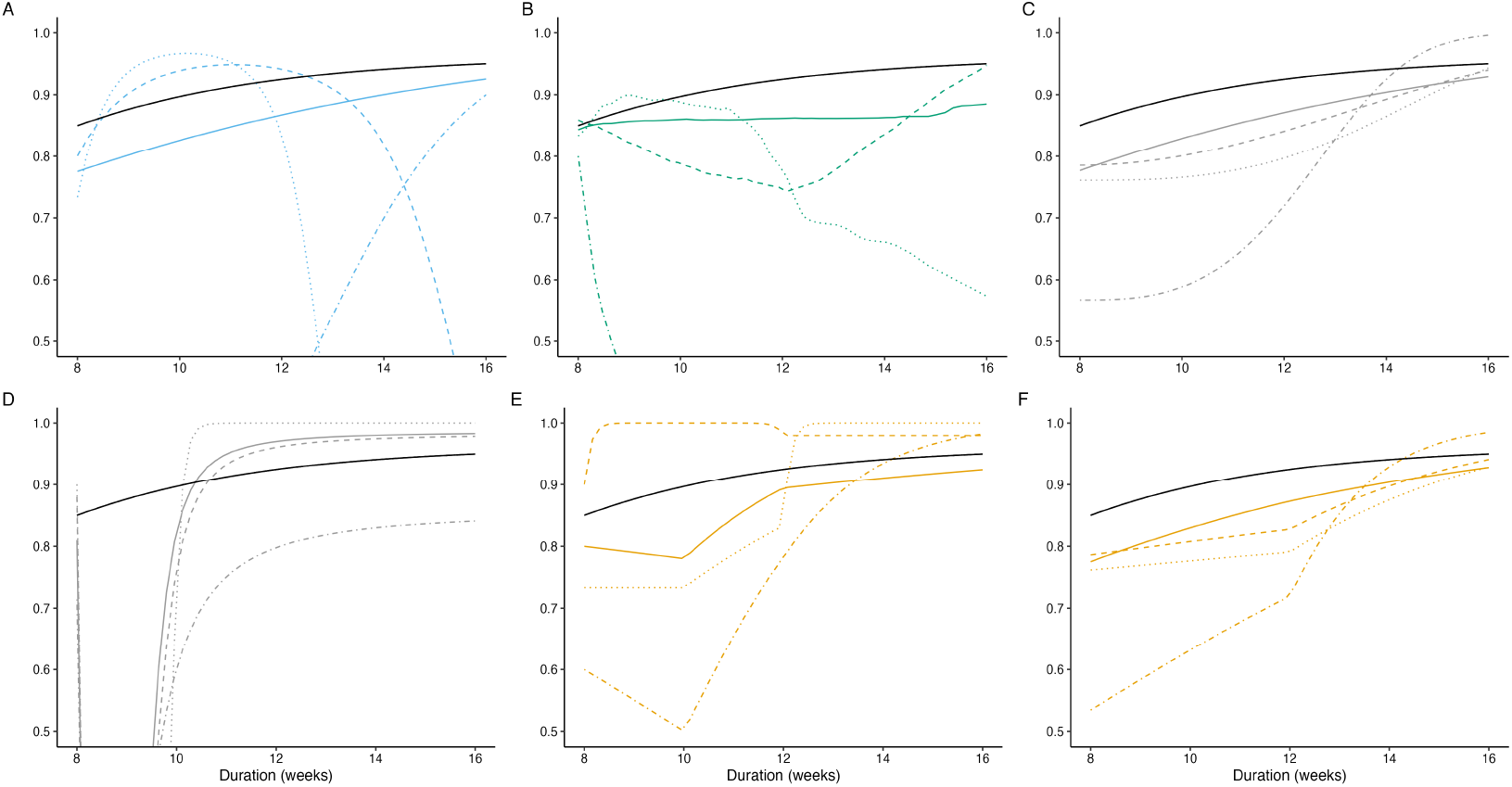
The estimated duration-response curves corresponding to the largest (worst) scaled area between the curves for sample sizes of 10 (dot-dashed), 30 (dotted), 50 (dashed), and 100 (solid) per duration when the true data-generating distribution is one of logistic growth (solid black line). A) MCP-Mod (select), B) MCP-Mod (average), C) FP1, D) FP2, E) LS2m, F) LS2e.

### 3.4 Performance: Estimation of Optimal Duration

Table 4 displays the true MED for each data-generating mechanism alongside the median estimated MED from applying each method to the 5,000 simulated datasets per data-generating mechanism with 30 participants per duration (*n*_*total*_ = 250). The Dunnett Test approach consistently failed to identify a MED less than the longest duration observed. Even at a sample size of 100 participants per duration, 82.2% of the estimates from the simulated datasets failed to identify a MED below 16 weeks (Supplemental Figure S2). All model-based methods, with the notable exception of MCP-Mod (average), display a tendency to underestimate the MED at this sample size. The magnitude of this underestimation is worth examining directly. For example, when the duration-response relationship is one of logistic growth, the median estimate of the minimum duration associated with a 90% probability of cure is 9.8 weeks whereas the truth is 10.2 weeks. Put another way, the estimated duration of approximately 9 weeks and 6 days is three days short of the true effective duration (10 weeks and 2 days). Is this cause for concern? Under this DGM, the true response rate at 9.8 weeks is 87.9% probability of cure, a full two percentage points lower than the targeted response rate. Careful consideration of acceptable error margins, or the use of a more conservative boundary such as the duration at which the estimated lower confidence bound exceeds 90%, will be critical in mitigating risk.

**Table 4:** The true minimum effective duration (MED) in weeks for each data-generating mechanism and the median (IQR) estimated MED from each method when applied across 5,000 simulated datasets with 30 participants per duration. Dunnett Test only shows results when the MED was estimated to be below 16 weeks (*<*5% of results).

|  | Logistic | Linear in Log-Odds | Linear |
| --- | --- | --- | --- |
| True MED | 10.2 | 11.1 | 12.0 |
| Estimated MED (IQR) |  |  |  |
| FP1 | 9.5 (8.2, 11.6) | 10.6 (8.2, 12.4) | 11.4 (8.3, 12.9) |
| FP2 | 9.6 (8.2, 10.9) | 10.1 (8.2, 11.8) | 10.6 (8.2, 12.2) |
| LS2e | 10.0 (8.7, 11.3) | 10.4 (8.7, 12.1) | 10.9 (8.7, 12.7) |
| LS2m | 9.3 (8.2, 10.8) | 10.1 (8.2, 11.6) | 10.1 (8.0, 12.1) |
| MCP-Mod (average) | 10.3 (8.3, 12.4) | 11.1 (8.7, 13.2) | 11.8 (8.5, 13.7) |
| MCP-Mod (select) | 9.8 (8.3, 11.6) | 10.6 (8.5, 12.2) | 11.1 (8.8, 12.9) |
| Dunnett Test (contiguous) | 14.0 (12.0, 14.0) | 14.0 (12.0, 14.0) | 14.0 (14.0, 14.0) |
| Dunnett Test (absolute) | 12.0 (10.0, 14.0) | 12.0 (10.0, 14.0) | 12.0 (10.0, 14.0) |

Less directly addressable via the previously proposed methods is the estimation risk associated with sampling error, which can be observed in the variation in estimated MEDs. The IQR for estimated MED for most model-based methods spans nearly 3-weeks, with 25% of the simulated data results returning estimated MEDs at or below 8 weeks. This behavior does improve as sample size increases (Supplemental Material, Table S3) and will benefit from the application of a more conservative boundary (e.g., using the duration whose lower confidence bound exceeds 90%), though the impact of random sampling error at standard Phase II sample sizes cannot be overstated.

The *POS*_MED_ performance results shown in Figure 6 aims to capture both the bias and variance in estimation by categorizing as “acceptable” any estimate of the MED within a prespecified interval around the true value, and “unacceptable” any estimate outside of this interval. The proportion of times a method returns an “acceptable” estimate can then be compared across methods and simulation settings. The first panel displays the results corresponding to “acceptable power”[19], or the probability that the estimated optimal duration will be shorter than the maximum observed but no shorter than the true optimal duration. This interval attempts to rule out the possibility of underestimating the duration required to achieve a 90% response rate. The second panel allows underestimation of the true optimal duration by up to 1 week, while also setting an expectation of no more than a 2 week overestimation of the true optimal duration. The third panel allows for underestimation of 2 weeks and expects an overestimation of no more than 4 weeks for estimation of the true optimal duration.

**Figure 6:**
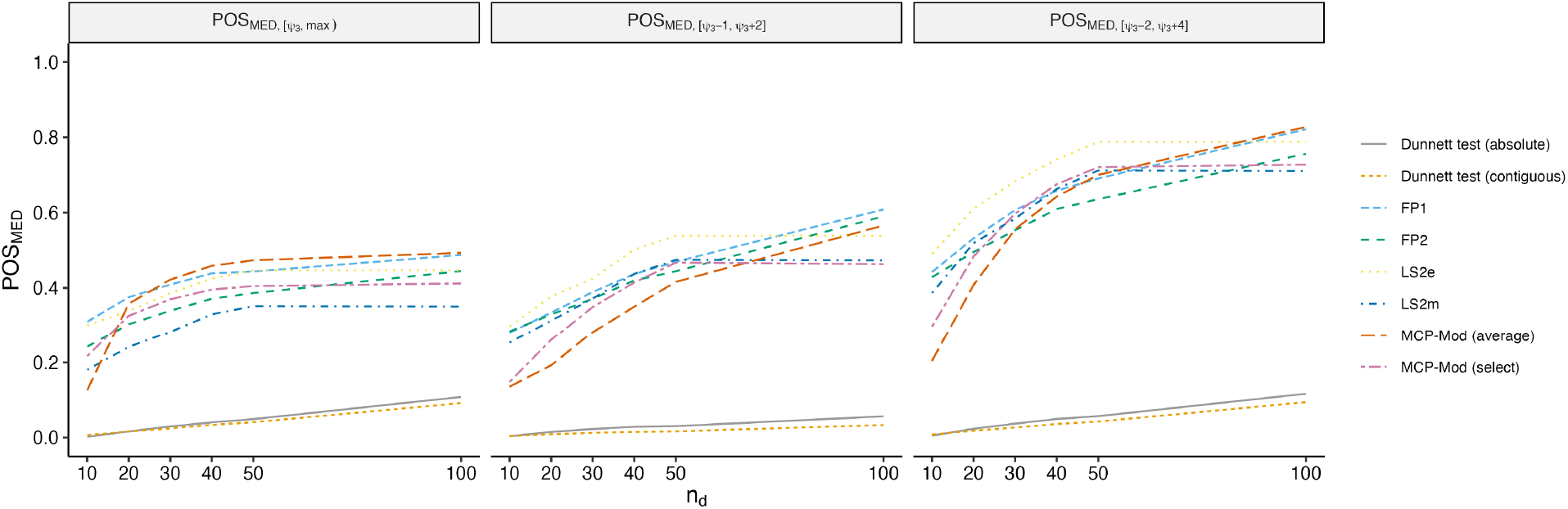
Estimated POS_MED_ for A) POS_MED,[0,max)_, B) POS_MED,[−1,+2]_, and C) POS_MED,[−2,+4]_ across 15,000 simulated datasets (5,000 simulated datasets for each DGM).

The improvement in POS_MED_ performance observable across the three panels for most of the model-based methods appears to arise primarily as a result of accepting slight underestimation of the optimal duration. The first panel (Figure 6: POS_MED,[*ψ*3, *max*]_) appears to contain the poorest proportion of acceptable results, despite defining acceptable as an estimated MED anywhere between the truth and the maximum duration observed (16 weeks). The second panel (Figure 6: POS_MED,[*ψ*3−1,*ψ*3 +2]_) returns what appear to be improved performance results despite a far stricter interval, only allowing overestimation of the true MED by 2 weeks yet allowing *underestimation* of up to one week from the true MED. The only method whose performance appears worse when comparing these two panels is the MCP-Mod (average) method, which appears to be the only method with a tendency to overestimate the MED. The last panel (Figure 6: POS_MED,[*ψ*3−2,*ψ*3 +4]_) has an upper bound on the acceptable region of estimated MEDs that is similar to the first, allowing overestimation by up to four weeks. However, the performance is strikingly high compared to that displayed in the first panel, which seems to suggest a tendency towards underestimation as this last panel categorizes underestimates of up to two weeks as acceptable. This is potentially concerning behavior worthy of further exploration. In the setting examined, there is very little difference in the true response rate at the true optimal duration and a duration of one or two weeks less, perhaps mitigating concern about the results of potentially under-prescribing treatment. However, if the true underlying duration-response curve has a more extreme shape and covers a wider range of response rates, then the risk associated with underestimation of the optimal duration will naturally increase.

## 4 Discussion

In this work we have adapted popular model-based dose-ranging techniques, including MCP-Mod and fractional polynomials, for the purposes of duration-ranging and have shown through simulation that such model-based methods outperform standard qualitative comparisons on every target, particularly in the constrained sample sizes of a Phase II trial. Though no method, model-based or qualitative, achieved high power at the lowest sample sizes for the simulation settings examined, FP1 showed the highest power in detecting a non-flat duration-response relationship. With respect to accurately reproducing the true duration-response curve, MCP-Mod and FP1 had the lowest average error across all sample sizes considered. As previously mentioned, this task is not feasible with the qualitative methods and therefore is a direct advantage of implementing a model-based duration estimation strategy. The linear spline approaches were relatively unreliable in replicating the duration-response curve, with knot specification playing a critical and overly influential role. Finally, as in the dosing literature, we observed the tendency to overestimate the optimal duration when standard qualitative comparisons were implemented for analysis. This effect is, in part, due to the inability to directly target the desired MED and instead having to apply a conservative testing framework. Model-based methods, on the other hand, were much more agile in identifying the MED, but not without concerns for underestimation. Such concerns could be mitigated upon the application of a more conservative threshold (e.g., estimating the MED based on an estimated lower confidence bound rather than the point estimate). However, the use of such an approach should take into account model-selection when estimating confidence intervals in the case of the fractional polynomial models. This is further described by Pham *et al* (forthcoming). FP2 appears to be particularly susceptible to underestimation of the MED at smaller sample sizes. In summary, it seems that MCP-Mod and FP1 would be reasonable approaches for future trials targeting the estimation of the optimal duration, with MCP-Mod using model-averaging at a slight advantage when more complex duration-response curves are expected.

In placebo-controlled dose-ranging, evidence of effect (*ψ*_1_) must typically be established in order to determine the optimal dose. In the duration-ranging setting where a known effective duration serves as the maximum duration under evaluation, establishment of a duration-response relationship is not a requirement and, in fact, its absence may be desirable in that it suggests the maximum duration may not be the optimal duration. In this sense, estimating the optimal duration is philosophically distinct from estimating optimal dose; hence, estimating the MED (*ψ*_3_) is not dependent on establishing evidence of effect (*ψ*_1_).

This work did not examine the impact of design parameters on performance, such as the number of durations observed, the spacing between durations, nor the (im)balance of sample size across durations. The work by [4] suggests that reasonable coverage of the duration-response curve, even if durations are non-equidistant and sample size is somewhat imbalanced, should result in similar results though, naturally, be subject to error when unanticipated non-linearity of the curve is not observed.

Future work is underway to incorporate patient characteristics and risk profiles into both the data-generating mechanism and the method of analysis. Information on the inclusion of covariates for the MCP-Mod procedure is available, but limited. Further development will be needed to inform the inclusion of covariate information in a duration-response setting. This work also does not address the setting where the true MED falls outside of the observation region, either at a lower duration than randomized to or, in a less likely scenario, at a higher duration than the maximum observed.

At the 2014 EMA/EFPIA workshop on dose finding, the gathered experts came to the clear and compelling agreement that “selection of dose for phase III is an estimation problem and should not be addressed via hypothesis testing” [27]. Duration ranging should equally receive such directional reframing. Randomized trial design must evolve to hone the ability to directly estimate the optimal duration. In this regard, designs such as MAMS-ROCI[19] and TB therapeutics trials such as SPECTRA-TB (ACTG A5414) and DRAMATIC (NCT03828201) lead the way toward a new era of treatment shortening efforts.

## Data Availability

All code necessary for data generation, analysis, and report compilation is publicly available in a GitHub repository maintained by the first author (https://github.com/sdufault15/tb-mcp-mod).

## Supplemental Material

### S1 Data-Generating Mechanisms

**Table S1:**
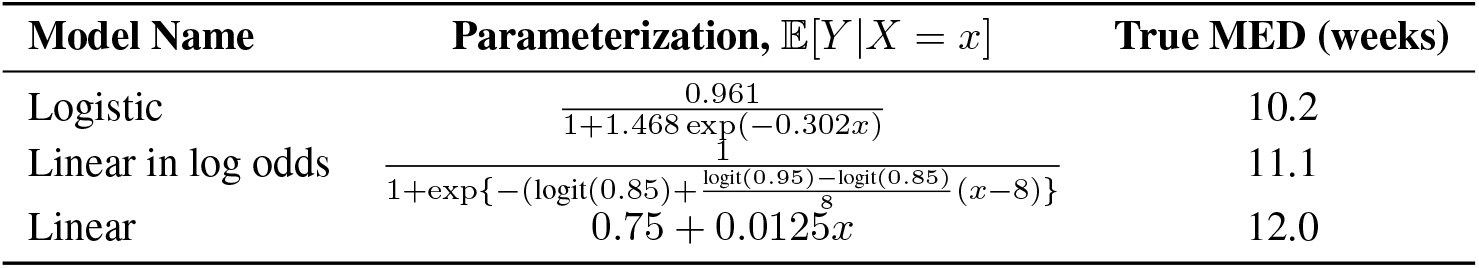
Parameterizations of the data-generating mechanisms used for the simulation study and the corresponding minimum effective duration (MED) associated with a 90% response rate.

### S2 Methods of Analysis: Parameterizations

#### MCP-Mod

Because the MCP-Mod procedure in R [21] expects

**Table S2:**
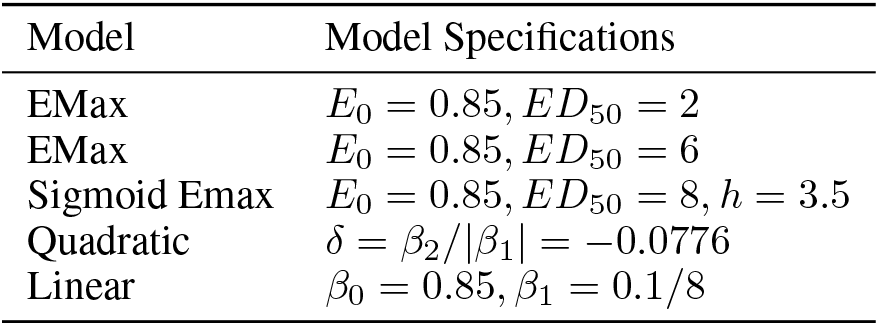
The models included in the MCP-Mod candidate library and their initial specifications.

**Figure S1:**
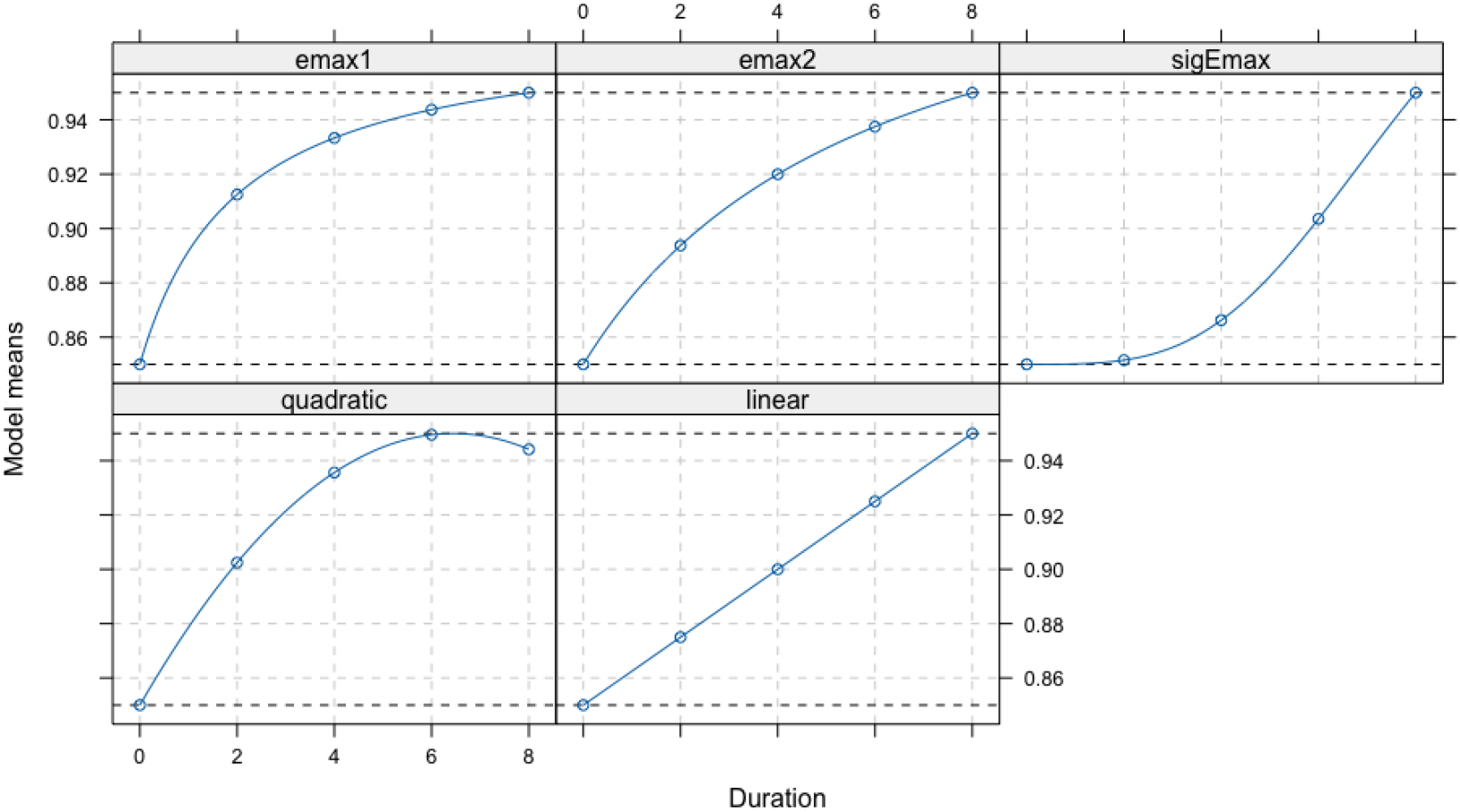
The shapes included in the MCP-Mod candidate library.

#### Fractional Polynomials

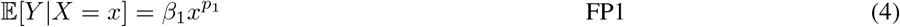

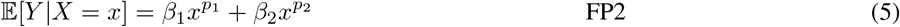

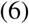

Where the powers *p*_1_, *p*_2_ are taken from the set *S* = {−2, −1, −0.5, 0, 0.5, 1, 2, 3 } and selected by the internal `gamlss` algorithm based on best fit for each simulated dataset.

#### Linear splines

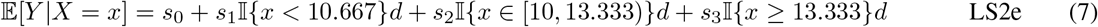

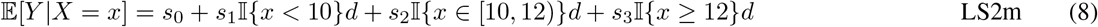

### S3 Additional Results

**Table S3:** The true minimum effective duration (MED) in weeks for each data-generating mechanism and the median (IQR) estimated MED from each method when applied across 5,000 simulated datasets with 100 participants per duration.

|  | Logistic | Linear in Log-Odds | Linear |
| --- | --- | --- | --- |
| True MED | 10.2 | 11.1 | 12.0 |
| LS2e | 12.1 (10.8, 12.9) | 10.9 (10.1, 12.2) | 10.3 (9.6, 10.9) |
| LS2m | 11.4 (10, 12.7) | 10.8 (9.5, 11.8) | 10.1 (9.3, 10.9) |
| MCP-Mod (average) | 11.9 (10.8, 12.9) | 11.1 (9.8, 12.1) | 10.1 (9.1, 11.1) |
| MCP-Mod (select) | 11.4 (9.6, 12.9) | 10.6 (9, 12.1) | 10 (8.7, 11.3) |
| FP1 | 12.1 (10.8, 12.9) | 11.1 (9.6, 12.1) | 10 (8.7, 11.1) |
| FP2 | 11.6 (10.4, 12.7) | 10.8 (9.3, 11.8) | 10.1 (8.2, 10.9) |
| Dunnett (absolute) | 14 (14, 14) | 14 (12, 14) | 14 (12, 14) |
| Dunnett (contiguous) | 14 (14, 14) | 14 (14, 14) | 14 (12, 14) |

**Figure S2:**
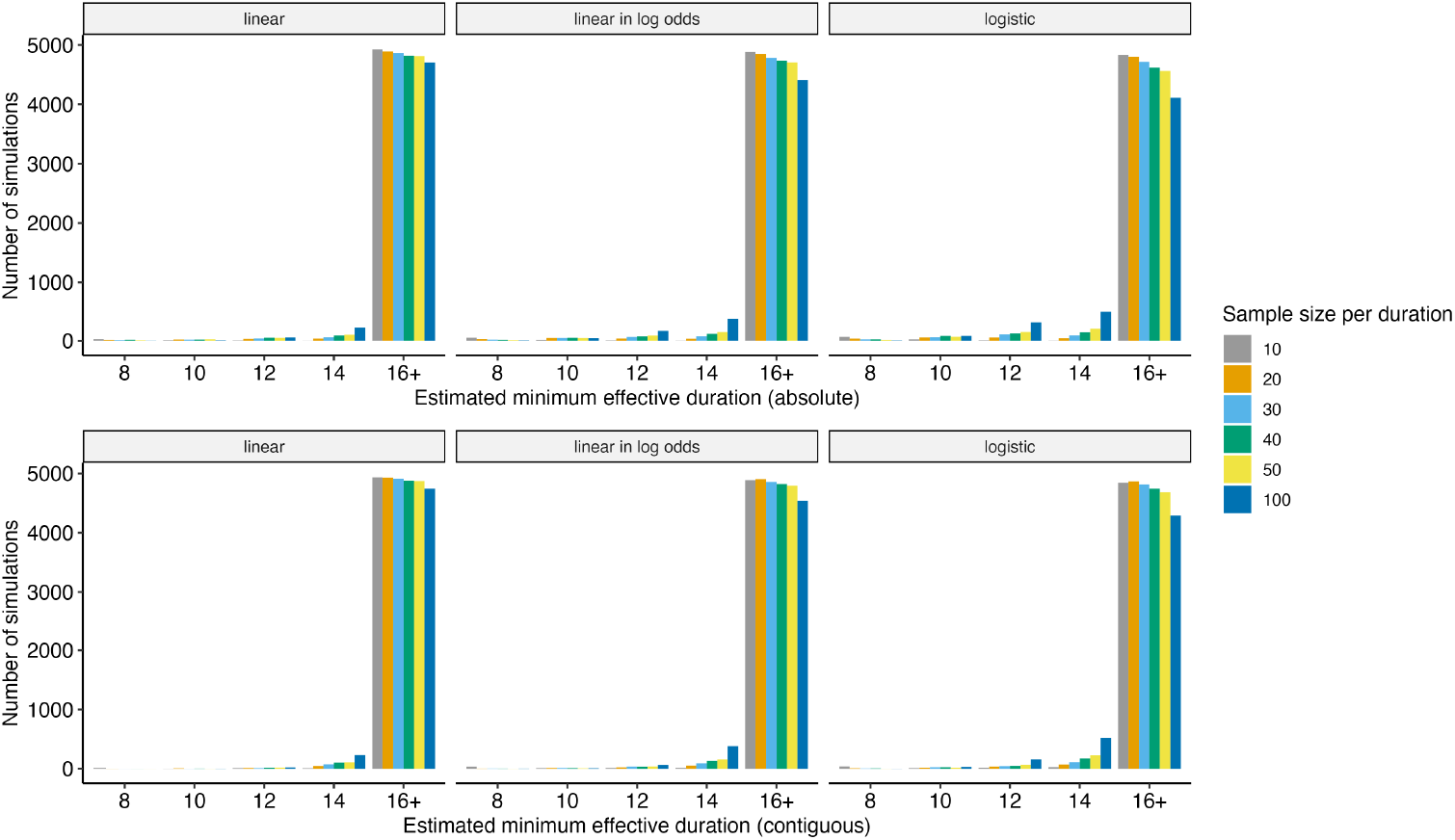
The estimated minimum effective durations from applying the Dunnett Test to 5,000 simulated datasets at each sample size.

**Figure S3:**
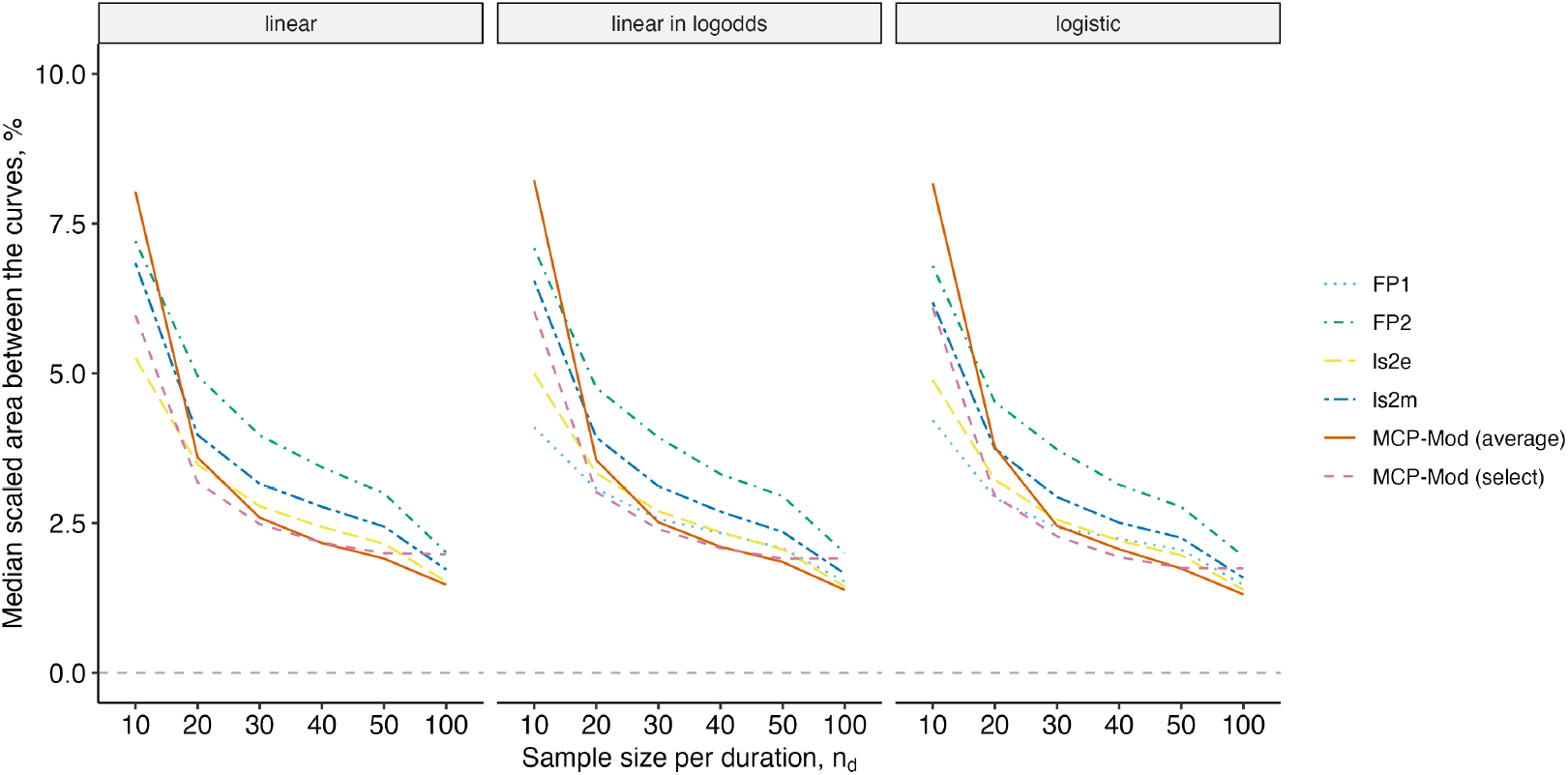
The median scaled area between the curves (%) from the estimated fits from 5,000 simulated datasets of each sample size (*n*_*d*_) for each data-generating mechanism and modeling method. A scaled area between the curves (%) of zero would indicate perfect fit.

